# Sleep duration and the risk of recurrent arrhythmias after catheter ablation of atrial fibrillation: the PREDIMAR study

**DOI:** 10.64898/2026.07.10.26357791

**Authors:** Jesús Díaz-Gutiérrez, María Teresa Barrio-López, Leticia Goni, Pablo Ramos, Luis Tercedor, José Luis Ibáñez, Gonzalo Barón-Esquivias, Eduardo Castellanos, Alicia Ibáñez-Criado, Rosa Macías-Ruiz, Ignacio García-Bolao, Miguel Ángel Martínez-González, Jesús Almendral, Miguel Ruiz-Canela

## Abstract

**Background:** Short and long sleep duration have been linked to atrial fibrillation (AF), but their influence on arrhythmia recurrence after catheter ablation is uncertain. We evaluated the association between nocturnal sleep duration and the risk of recurrent arrhythmias in patients undergoing catheter ablation for AF in the PREDIMAR trial.

**Methods:** The PREDIMAR study is a multicentre, randomized, controlled, single-blind trial evaluating a Mediterranean diet enriched with extra-virgin olive oil for preventing arrhythmia recurrence after catheter ablation for AF. Nocturnal sleep duration was categorized as adequate (6–8 h/day) or inadequate (<6 h/day or >8 h/day). Multivariable Cox regression models estimated the association between sleep duration and the risk of recurrent atrial flutter (AFL) or AF.

**Results:** Among 720 participants, we observed 226 incident cases of AF relapse and 107 cases of AFL. Inadequate nocturnal sleep duration was associated with a significantly higher risk of AFL recurrence compared with adequate sleep (adjusted HR = 1.87; 95% CI 1.18–2.96). No significant association was observed for AF recurrence (HR = 0.99; 95% CI 0.70–1.41). The association with AFL recurrence was particularly evident in patients with persistent AF at baseline before ablation (adjusted HR = 3.42; 95% CI 1.47–7.97), whereas no significant relationship was observed in those with baseline paroxysmal AF.

**Conclusions:** Inadequate nocturnal sleep duration (<6 h/day or >8 h/day) may increase the risk of AFL recurrence following AF ablation. These findings highlight the relevance of sleep habits as a modifiable behavioural factor potentially influencing post-ablation outcomes.

**Registration:** https://www.clinicaltrials.gov; Unique Identifier: NCT03053843.

**Clinical Perspective:** *What Is New?:*

- In patients undergoing catheter ablation for atrial fibrillation, inadequate nocturnal sleep duration (<6 or >8 h/day) was independently associated with a nearly two-fold higher risk of atrial flutter recurrence, with no significant association for atrial fibrillation recurrence.
- The association with atrial flutter recurrence was especially pronounced among patients with persistent atrial fibrillation before ablation.

*What Are the Clinical Implications?:*

- Assessing and optimizing sleep duration may represent a modifiable behavioural target to improve arrhythmia-free outcomes after atrial fibrillation ablation, particularly in patients with persistent atrial fibrillation.

## INTRODUCTION

Atrial fibrillation (AF) is the most common sustained cardiac arrhythmia and is associated with considerable morbidity and mortality^1^. Catheter ablation has become an effective therapeutic option for rhythm control in patients with AF; however, recurrence rates remain substantial despite technological advances and optimized medical therapy^2^. Identifying modifiable behavioural factors that may influence post-ablation outcomes could improve long-term rhythm maintenance and reduce disease burden. In this context, lifestyle-related determinants such as sleep warrant particular attention.

Sleep is a key determinant of cardiovascular health. The American Heart Association recently incorporated sleep into its eight essential metrics of cardiovascular health^3^. A growing body of evidence suggests that sleep habits influence the development of AF and other cardiovascular conditions. Large prospective cohort studies have shown that both short and long sleep duration are associated with an increased risk of cardiovascular disease, including coronary artery disease, stroke, and AF^4–6^. A recent meta-analysis of eight observational studies reported an 18% higher risk of incident AF among individuals sleeping less than 6 hours per night, whereas long sleep duration was not significantly associated with AF risk^7^.

Whether sleep duration also affects AF recurrence after catheter ablation remains insufficiently understood. In a recent study of patients undergoing catheter ablation, Cang *et al.* reported that an i, including short or long sleep duration, was independently associated with an increased risk of AF recurrence^8^. Other studies have similarly suggested that both insufficient and excessive sleep may influence post-ablation outcomes, although findings have not been entirely consistent^9,10^.

Given the accumulating evidence linking inadequate sleep duration to incident AF, clarifying its potential influence on post-ablation arrhythmia recurrence is of clinical and preventive relevance. In the present analysis of the PREDIMAR trial, we investigated the association between sleep duration and the risk of recurrent atrial arrhythmias following AF ablation.

## METHODS

### Study Population

The PREDIMAR study is a multicentre, randomized, controlled, single-blind clinical trial evaluating whether a Mediterranean diet enriched with extra-virgin olive oil can reduce the recurrence of atrial tachyarrhythmias after catheter ablation for AF. Following successful ablation, all eligible patients in sinus rhythm who provided written informed consent were randomly assigned in a 1:1 ratio to either the intervention group (Mediterranean diet supplemented with extra-virgin olive oil) or the control group (usual clinical care). Clinical staff involved in patient management were blinded to treatment allocation. Detailed information on the study design and protocol has been reported elsewhere^11^.

Eligible participants had (i) symptomatic paroxysmal AF before ablation, defined as more than one symptomatic episode in the previous year with at least one documented event, or (ii) symptomatic persistent AF before ablation. Recruitment took place in four Spanish hospitals: Hospital Montepríncipe (Madrid), Clínica Universidad de Navarra (Pamplona), Hospital Virgen de las Nieves (Granada), and Hospital General Universitario de Alicante (Alicante). Between March 2017 and January 2020, a total of 720 participants were enrolled, randomized, and began the intervention.

The trial was registered at ClinicalTrials.gov (identifier NCT03053843). The protocol was approved by the Research Ethics Committees of all participating centres, and all participants signed informed consent forms after receiving written and verbal explanations of the study procedures.

### Catheter ablation procedure

Ablation procedures were performed under general anaesthesia, predominantly using radiofrequency energy. Two catheters were advanced into the left atrium through a single transseptal puncture or a patent foramen ovale: a 20-pole circular mapping catheter (Advisor, Abbott) and a contact-force sensing irrigated ablation catheter (Tacticath Quartz, Abbott). Radiofrequency energy was delivered at 30–50 W with a temperature limit of 43–46 °C and a target contact force of 10–40 g. In all procedures, the ablation catheter was manipulated using a deflectable sheath (Agilis, Abbott). Pulmonary vein isolation was verified by confirming bidirectional conduction block after a 30-minute waiting period, followed by adenosine administration to identify dormant conduction. Intravenous heparin was administered to maintain an activated clotting time ≥350 s.

### Assessment of sleep duration, lifestyle habits and other covariables

Lifestyle data were obtained at baseline (immediately after a successful ablation procedure) and at 12- and 24-month follow-up interviews conducted by trained dietitians via telephone. The questionnaires covered lifestyle-related data including sleep, dietary habits, physical activity, and sedentary behaviour.

Average nocturnal sleep and daytime nap duration were derived from the questions: “How many hours per day do you sleep at night?” and “How many hours per day do you take a midday nap?”. Possible responses ranged from “never” to categories spanning “<30 min/day” to “>9 h/day”, separately for weekdays and weekends. Weighted averages were computed (5 × weekdays + 2 × weekends / 7). Nocturnal sleep duration was initially analysed as a discrete variable and later categorized as adequate (6–8 h/day) or inadequate (<6 h/day or >8 h/day)^5^. Daytime nap duration was classified into three groups: <30 min/day, ≥30 min/day, and no nap^12^.

Adherence to the traditional Mediterranean diet was assessed using the validated 14-item Mediterranean Diet Adherence Screener (MEDAS; range 0–14)^13^. Dietary intake was evaluated through a 147-item semi-quantitative food frequency questionnaire (FFQ) previously validated for the Spanish population^14^. Physical activity was quantified using a previously validated questionnaire for Spanish adults^15^. A physical activity score (range 0–8) was estimated to capture the overall pattern of physical activity and avoidance of sedentary behaviours (time watching television and sitting time)^16^.

Baseline data also included sociodemographic, clinical, and anthropometric characteristics. We assessed the prevalence of cardiovascular risk factors (smoking, hypertension, dyslipidaemia, and diabetes), history of cardiovascular disease (heart failure, cardiomyopathy, or valvular heart disease), antiarrhythmic drug use, time from the first diagnosis of AF to the ablation procedure, any other previous ablation, presence of sleep apnoea, and working hours.

Left atrial (LA) size was assessed using echocardiographic diameter, area, and volume, each categorized into four severity groups. Echocardiographic LA enlargement was defined as the highest severity category across these measures. CT-derived LA size was similarly classified into four levels, and a composite two-category variable for overall LA enlargement (none/mild and moderate/severe) was created by prioritizing CT data when available^17^.

### Follow-up and outcome ascertainment

The active dietary intervention lasted two years, with clinical visits scheduled at 3, 6, 12, 18, and 24 months after ablation. At each visit, a 12-lead ECG and 24-hour to 7-day Holter monitoring were performed. In addition, participants received a portable electrocardiographic device (*Kardia Mobile*) between month 3 and 18 of follow-up and were instructed to record a one-minute ECG weekly and whenever symptoms occurred. Additional sources of rhythm documentation included recordings from cardiac implantable electronic devices, when present, and 12-lead ECG obtained during emergency department visits for arrhythmia-related symptoms. Recurrent arrhythmia was defined as any documented AF or AFL lasting ≥30 seconds and occurring after a three-month blanking period until month 18, the period when *Kardia* devices were systematically used (>90%) to detect endpoints.

### Statistical Analysis

Baseline characteristics were summarized as percentages or means and standard deviations across sleep duration categories.

We used multivariable Cox regression models to assess the association between nocturnal sleep duration (two categories) with risk of recurrent AF or AFL. Hazard Ratios (HR) and their 95% Confidence Intervals (95% CI) were estimated, with sleeping 6-8 hours/day as reference category. Person-months of follow-up were calculated for each participant from randomization to the date of the documented arrhythmia recurrence, or the date of the 18-month visit, whichever occurred first.

Four multivariable models were used. Model 1 was adjusted for age (continuous), sex (male or female), intervention group (intervention or control), and recruitment center (four nodes); Model 2 with further adjustment for personal history of cardiovascular disease (i.e. heart failure, cardiomyopathy, or valvular heart disease; yes/no), hypertension (yes/no), diabetes (yes/no), hypercholesterolemia (yes/no), use of antiarrhythmic drugs (yes/no), any other previous ablation (yes/no), time from the first diagnosis of AF to the ablation procedure (continuous), and left atrial enlargement (none/mild or moderate/severe); Model 3 with further adjustment for overweight and obesity (yes/no), smoking habit (active smoker, former smoker, or never smoker), physical activity (tertiles), sleep apnoea (yes/no), coffee intake (<1 cup/day, 1 cup/day, or >2-3 cups/day), caffeine intake (tertiles), tea intake (continuous), alcohol binge drinking (more than 5 alcoholic drinks at any time; yes/no), Mediterranean diet adherence score (0 to 14 points; tertiles), working hours (not working, ≤34 h/week, 35-39 h/week, 40-44 h/week, ≥45 h/week), and educational level (university studies; yes/no); and Model 4 (main analysis model), which additionally adjusted for daytime nap duration (no nap, <30 min/day, and ≥30 min/day).

Nelson-Aalen plots were used to show recurrent AFL incidence according to sleep duration (6-8 h/day vs. <6 h/day or >8 h/day). The incidence curves were adjusted for potential confounding using the inverse probability weighting method.

We further examined potential non-linear associations by modelling sleep duration using restricted cubic splines with three knots set at the 10th, 50th, and 90th percentiles (5, 7, and 8 h/day, respectively), adjusted according to Model 4.

For subgroup analyses, we stratified participants by the pre-ablation temporal pattern of AF (paroxysmal vs. persistent arrhythmia), body mass index (<25 kg/m^2^ vs. ≥25 kg/m^2^), sex (male vs. female), prevalent cardiovascular disease (presence vs. absence of heart failure, cardiomyopathy, or valvular heart disease), and daytime nap sleeping (yes vs. no). These analyses aimed to assess whether the association between sleep duration and recurrent AFL risk differed across relevant subgroups. Effect modification was assessed through the likelihood ratio test comparing models with and without the interaction term.

To evaluate the consistency of our findings, we performed several sensitivity analyses: 1) excluding participants younger than 45 years, 2) excluding those with prevalent cardiovascular risk factors (hypertension, diabetes, and dyslipidaemia), 3) excluding participants with prevalent cardiovascular disease (heart failure, myocardiopathy, or valvular heart disease), 4) excluding participants with prevalent sleep apnoea, 5) redefining adequate sleep duration as >6 hours/day, and 6) redefining adequate sleep duration as 7-8 hours/day.

All reported p values were two-sided, and statistical significance was set at p<0.05. Statistical analyses were performed using STATA/SE 17.0 (StataCorp, College Station, TX, US).

## RESULTS

### Baseline Characteristics

Baseline characteristics according to sleep duration categories are summarized in Table 1. Participants with inadequate sleep duration (<6 h/day or >8 h/day) were more frequently women, former smokers, not currently employed, and without higher education studies. They also had a slightly higher prevalence of obstructive sleep apnoea and reported lower coffee consumption. No clinically meaningful differences were observed across categories in cardiovascular risk factors, prevalent cardiovascular disease, physical activity levels, daytime nap duration, or other arrhythmia-related clinical features. Participants were followed-up for a median of 18 months, during which 107 recurrent AFL cases and 226 recurrent AF events were documented.

**Table 1.**
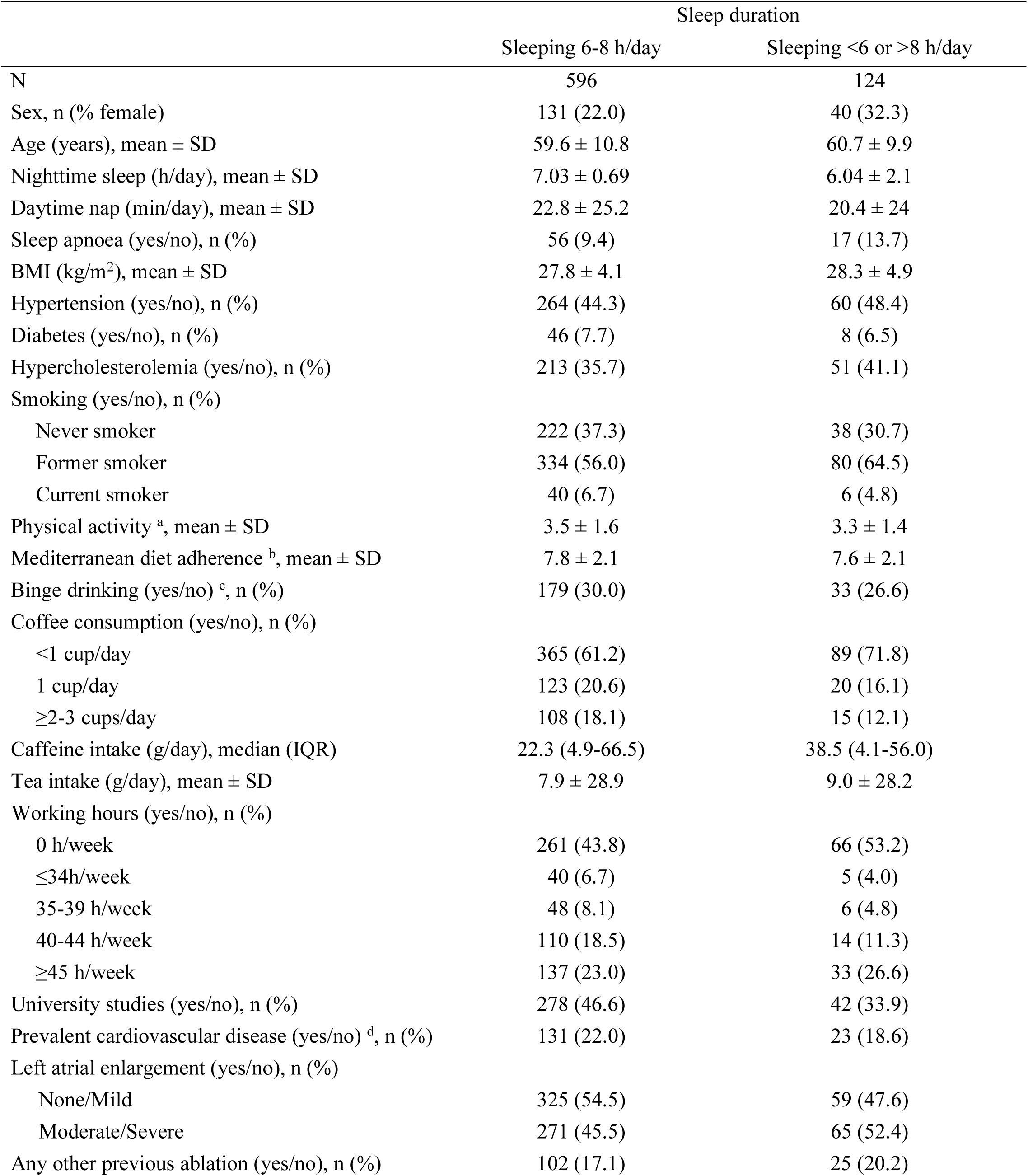
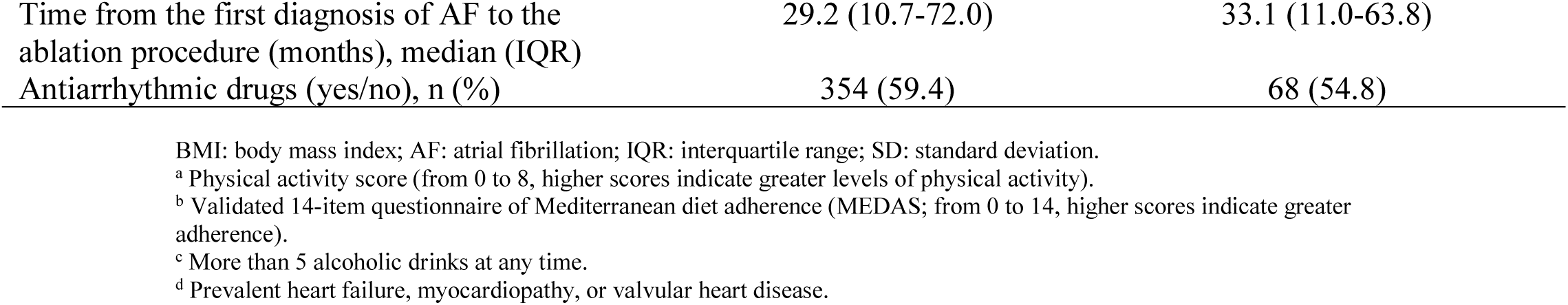
Baseline characteristics of participants according to night sleep time in PREDIMAR study.

### Sleep duration and risk of arrhythmia recurrence

As shown in Figure 1, participants with inadequate nocturnal sleep duration (<6 h/day or >8 h/day) had a 87% higher risk of AFL recurrence compared with those reporting adequate sleep (6–8 h/day) (adjusted HR = 1.87; 95% CI 1.18–2.96). By contrast, no significant association was observed for AF recurrence (adjusted HR = 0.99; 95% CI 0.70–1.41) (Table 2).

**Figure 1.**
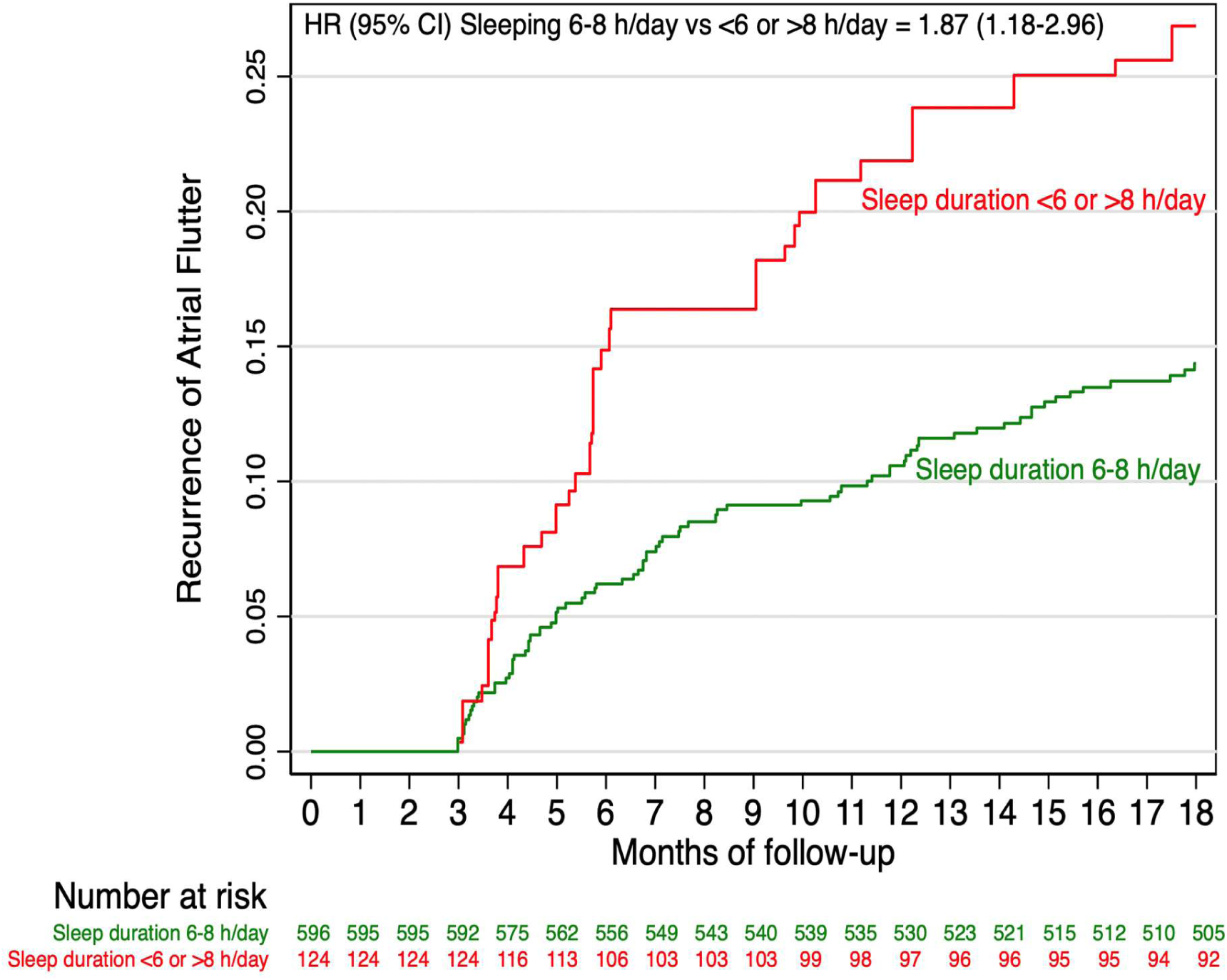
Nelson−Aalen survival plot for incident atrial flutter recurrence during follow-up according to categories of sleep duration. Footnote: Cumulative incidence curves are adjusted by inverse-probability weighting; numbers at risk are unweighted.

**Table 2.** Hazard Ratio and 95% Confidence Interval of arrhythmia recurrence according to nighttime sleep duration in PREDIMAR study after 18-month follow-up.

|  | Sleep duration |  |
| --- | --- | --- |
|  | Sleeping 6-8 h/day | Sleeping <6 or >8 h/day |
| **Atrial Flutter recurrence** |  |  |
| Participants, n | 596 | 124 |
| Cases/person-months, n/N | 78/322.6 | 29/62.1 |
| Model 1 <sup>a</sup> | 1 (Ref.) | 1.80 (1.16-2.78) |
| Model 2 <sup>b</sup> | 1 (Ref.) | 1.86 (1.20-2.90) |
| Model 3 <sup>c</sup> | 1 (Ref.) | 1.88 (1.19-2.97) |
| Model 4 <sup>d</sup> | 1 (Ref.) | 1.87 (1.18-2.96) |
| **Atrial Fibrillation recurrence** |  |  |
| Participants, n | 596 | 124 |
| Cases/person-months, n/N | 186/267.8 | 40/53.5 |
| Model 1 <sup>a</sup> | 1 (Ref.) | 1.00 (0.71-1.41) |
| Model 2 <sup>b</sup> | 1 (Ref.) | 0.97 (0.69-1.37) |
| Model 3 <sup>c</sup> | 1 (Ref.) | 0.99 (0.70-1.41) |
| Model 4 <sup>d</sup> | 1 (Ref.) | 0.99 (0.70-1.41) |

<sup>a</sup>Model 1: age, sex, intervention group, and recruitment center.
<sup>b</sup>Model 2: Model 1 + prevalent cardiovascular disease (heart failure, cardiomyopathy, and valvular heart disease), left atrial enlargement, antiarrhythmic drugs, any other previous ablation, time from the first diagnosis of AF to the ablation procedure, hypertension, diabetes, and hypercholesterolemia.
<sup>c</sup>Model 3: Model 2 + overweight/obesity, smoking, working hours, educational level, physical activity, sleep apnea, coffee intake, caffeine intake, tea intake, alcohol binge drinking, and Mediterranean diet adherence.
<sup>d</sup>Model 4: Model 3 + daytime nap duration.

The restricted cubic spline analysis illustrated a nonlinear relationship between sleep duration and the risk of AFL recurrence, with the lowest risk observed at approximately 7 hours of night sleep (Figure 2).

**Figure 2.**
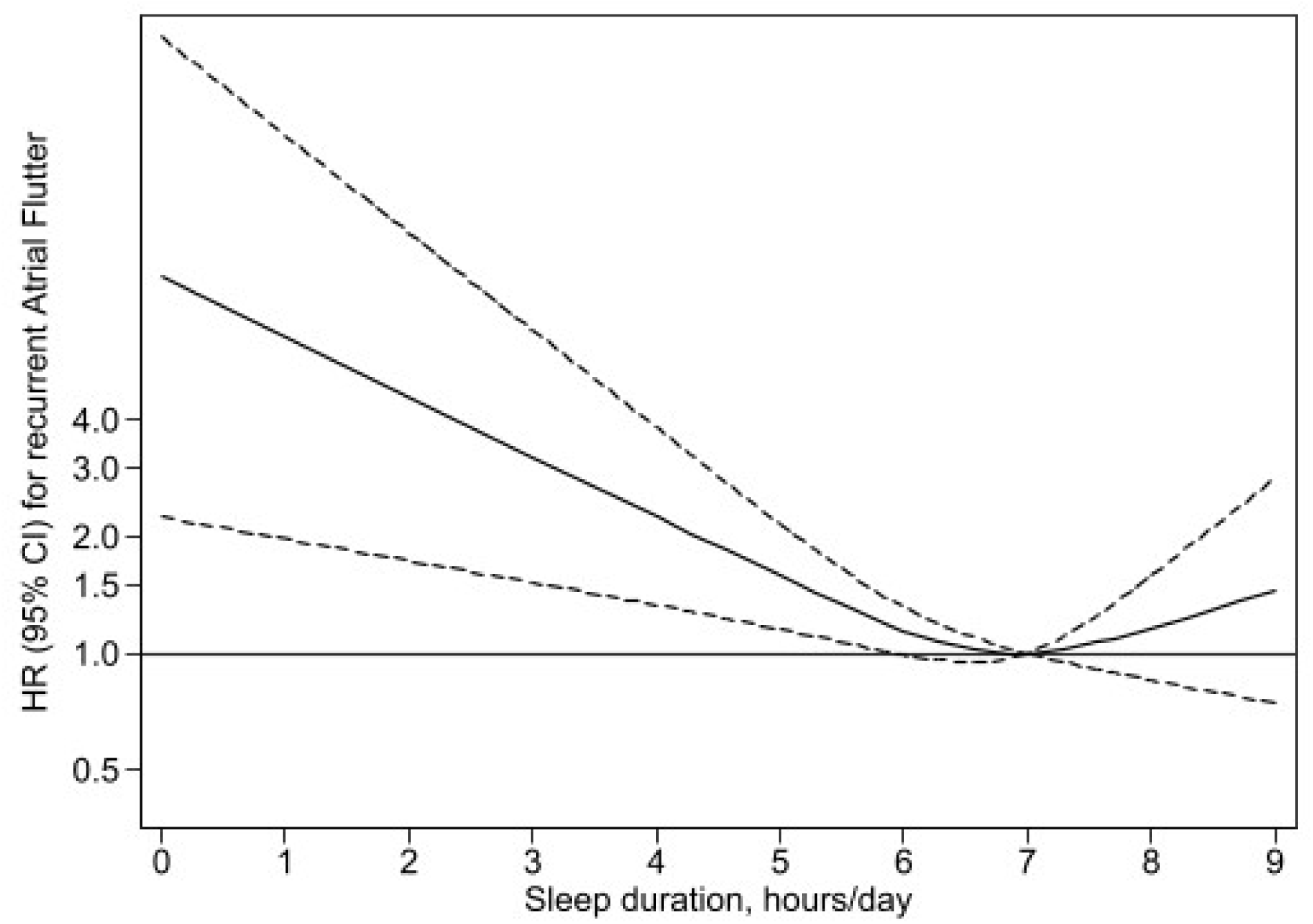
Restricted cubic spline regression for the association between sleep duration and the risk of incident atrial flutter recurrence.

Multiple sensitivity analyses confirmed the robustness of the association between sleep duration and the risk of AFL recurrence (Table 3). Notably, when adequate sleep was redefined as >6 h/day compared with <6 h/day, the association persisted with a slightly stronger effect estimate. Further adjustment for daytime nap duration (Supplementary Table 1) did not materially change the results, and no association was observed between daytime nap and AFL recurrence risk, suggesting that daytime sleep had little influence on the observed association.

**Table 3.** Sensitivity analyses for Atrial Flutter recurrence. PREDIMAR study.

|  | Adequate sleep duration<br>Sleeping 6-8 h/day |  | Inadequate sleep duration<br>Sleeping <6 or >8 h/day |  |
| --- | --- | --- | --- | --- |
|  | Cases/<br>Participants | HR<br>(95% CI) | Cases/<br>Participants | HR<br>(95% CI) <sup>a</sup> |
| Overall | 78/596 | 1 (Ref.) | 29/124 | 1.87 (1.18-2.96) |
| Only >45 years | 76/532 | 1 (Ref.) | 24/113 | 1.55 (0.94-2.54) |
| Only without cardiovascular risk factors <sup>b</sup> | 29/250 | 1 (Ref.) | 11/42 | 2.78 (1.11-6.94) |
| Only without prevalent cardiovascular disease <sup>c</sup> | 59/465 | 1 (Ref.) | 22/101 | 1.63 (0.96-2.75) |
| Only without sleep apnoea | 73/540 | 1 (Ref.) | 23/107 | 1.62 (0.97-2.71) |
| Recategorizing adequate sleep duration to >6 hours/day | 85/636 | 1 (Ref.) | 22/84 | 2.20 (1.34-3.62) |
| Recategorizing adequate sleep duration to 7-8 hours/day | 54/423 | 1 (Ref.) | 53/297 | 1.38 (0.92-2.08) |

- a. Adjusted for: age, sex, intervention group, recruitment center, prevalent cardiovascular disease (heart failure, cardiomyopathy, and valvular heart disease), left atrial enlargement, antiarrhythmic drugs, any other previous ablation, time from the first diagnosis of AF to the ablation procedure, hypertension, diabetes, hypercholesterolemia, overweight/obesity, smoking, working hours, educational level, physical activity, sleep apnea, coffee intake, caffeine intake, tea intake, alcohol binge drinking, Mediterranean diet adherence, and daytime nap duration. - b. Diabetes, hypertension, dyslipidemia. - c. Heart failure, cardiomyopathy, or valvular heart disease.

### Sleep duration and temporal pattern of atrial fibrillation

When stratified by the temporal pattern of arrhythmia, we observed a significant interaction on the multiplicative scale between sleep duration and AFL recurrence (likelihood ratio; p for the product term = 0.03). Among participants with pre-ablation paroxysmal AF, inadequate nocturnal sleep (<6 h/day or >8 h/day) was not associated with AFL recurrence (adjusted HR = 1.73; 95% CI 0.80–3.73). By contrast, in those with persistent AF, inadequate sleep duration was associated with a higher risk of AFL recurrence (adjusted HR = 3.42; 95% CI 1.47–7.97) (Table 4).

**Table 4.**
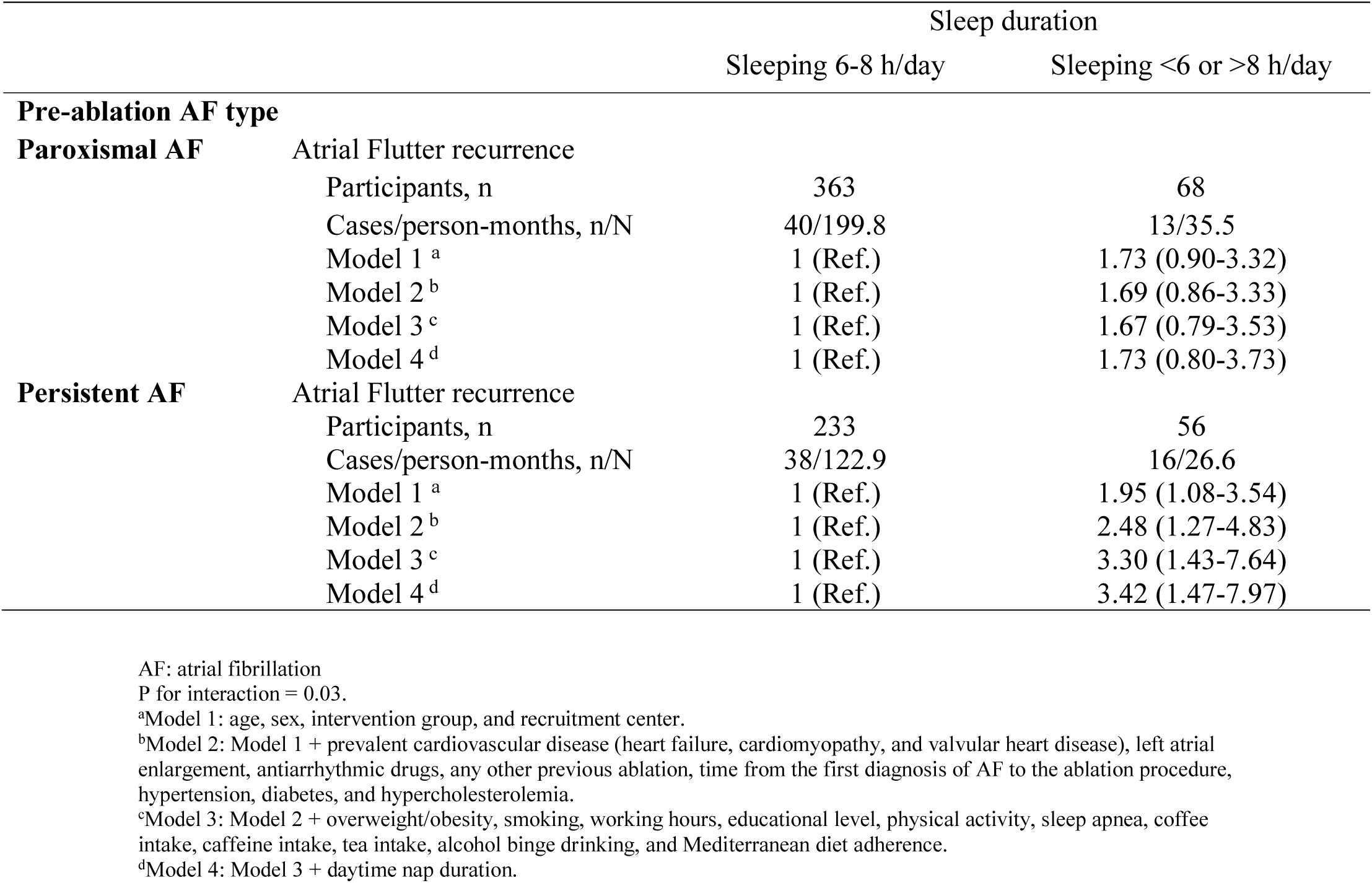
Hazard Ratio and 95% Confidence Interval of Atrial Flutter recurrence according to nighttime sleep duration in PREDIMAR study after 18-month follow-up and stratified by the pre-ablation arrhythmia temporal pattern.

In additional subgroup analysis, there was no heterogeneous effect on the risk of AFL recurrence depending on body mass index (p for interaction = 0.39), sex (p for interaction = 0.39), prevalent cardiovascular disease (p for interaction = 0.34), or daytime nap sleeping (p for interaction = 0.60).

## DISCUSSION

In this prospective analysis of patients with AF undergoing catheter ablation within the PREDIMAR trial, inadequate nocturnal sleep duration (<6 h/day or >8 h/day) was associated with a higher risk of AFL recurrence. This relationship remained robust across multiple sensitivity analyses and showed a nonlinear pattern, with the lowest risk observed at approximately 7 hours of night sleep. When stratified by the temporal pattern of arrhythmia, the association was particularly evident among patients with pre-ablation persistent AF, whereas no significant relationship was observed in those with pre-ablation paroxysmal AF. No association was observed between night sleep and AF recurrence.

Our findings for AFL need to be interpreted in the context of previous evidence suggesting a relationship between sleep duration and risk of incident atrial tachyarrhythmias. Population-based cohorts, including the UK Biobank, and large Chinese and U.S. cohorts, have consistently reported U-shaped associations between sleep duration and the incidence of AF, with both short and long sleep linked to higher AF risk^18–20^. These observations have been supported by Mendelian randomization analyses suggesting a potential causal effect of short sleep duration on AF development^4,21^. A recent meta-analysis of eight observational studies further confirmed a higher risk of AF among individuals sleeping less than 6 hours per night, while longer sleep durations showed no clear association^7^.

However, these studies mainly addressed the primary prevention of AF in the general population rather than arrhythmia recurrence after catheter ablation. Evidence in the secondary prevention setting remains limited. In a cohort study by Cang et al., an unhealthy sleep pattern, including short or long sleep duration, was independently associated with an increased risk of AF recurrence after ablation^8^. Similarly, in a study of patients undergoing catheter ablation for persistent AF, habitual long objective sleep duration was identified as a predictor of AF recurrence^9^. Conversely, another investigation reported that shorter sleep duration (≤7 h/day) was associated with higher recurrence risk, whereas longer sleep duration showed no significant association^10^.

Our study expands on these findings by incorporating repeated rhythm monitoring and comprehensive adjustment for clinical and lifestyle factors in a well-characterized Mediterranean cohort. These results reinforce the notion that inadequate sleep may play a role not only in the onset but also in the recurrence of atrial arrhythmias. In contrast to previous reports, we did not observe an association between sleep duration and AF recurrence. Rather, the association in our cohort was restricted to AFL recurrence, particularly among patients with persistent AF. This distinction may reflect differences in study populations, arrhythmia phenotypes, follow-up strategies, or the underlying mechanisms driving AF versus other atrial tachyarrhythmias.

In our study, the association between inadequate sleep duration and AFL recurrence appeared to differ according to the temporal pattern of AF. The relationship was stronger among patients with persistent AF, whereas no significant association was observed in those with paroxysmal AF. These findings suggest that the detrimental effects of inadequate sleep may be more pronounced in individuals with a more advanced arrhythmic substrate, in whom structural and electrical remodelling are already established^22^.

A key question raised by our findings is why inadequate sleep duration was associated with AFL, but not AF, recurrence, given that the autonomic, circadian, and inflammatory pathways implicated in sleep-related arrhythmogenesis might be expected to influence both. One plausible explanation lies in the distinct substrates underlying each arrhythmia. Whereas AF recurrence after pulmonary vein isolation is frequently driven by triggers and reconnection at the vein level^23^, macro-reentrant atrial tachyarrhythmias such as typical flutter depend on a more advanced and organized atrial substrate, with established structural and electrical remodelling that sustains stable reentrant circuits^24,25^. Inadequate sleep, through sustained sympathetic activation, reduced vagal tone, and circadian disruption, may act preferentially on this remodelled substrate, promoting the organization of reentry rather than the trigger-dependent mechanisms characteristic of AF. This interpretation is consistent with our observation that the association was confined to patients with persistent AF at baseline, in whom the atrial substrate is typically more remodelled, and absent in those with paroxysmal AF. We propose this as a biologically coherent hypothesis rather than a definitive mechanism, and one that warrants confirmation in studies designed with objective sleep assessment and detailed characterization of the flutter circuit.

These mechanisms can be understood at the level of the substrate on which inadequate sleep acts. Beyond well-established structural determinants of left atrial fibrosis, increasing attention has been directed towards the role of autonomic activity and circadian regulation in AF pathophysiology. Variations in circadian rhythm have been linked to both the initiation and maintenance of AF^26^, and sleep stages may influence arrhythmogenesis through their effect on autonomic cardiac signalling, which plays a critical role in AF development^27^.

Insufficient sleep enhances sympathetic activation and reduces vagal tone, leading to greater heart rate variability and increased atrial excitability^28^. Experimental models indicate that sleep restriction can elevate circulating catecholamine levels and may modify the atrial effective refractory period, thereby facilitating ectopic activity and re-entry circuits^29^. Sleep restriction has also been associated with higher levels of inflammatory cytokines and increased cortisol secretion through stimulation of the hypothalamic–pituitary–adrenal axis^30,31^. Together, these responses may promote endothelial dysfunction, oxidative stress, and atrial fibrosis, providing a substrate for electrical and structural remodelling that facilitates arrhythmia recurrence^22^.

Although the precise mechanisms linking sleep duration to post-ablation outcomes remain to be elucidated, the interplay of neuroendocrine imbalance, inflammatory activation, and circadian disruption provides a biologically plausible framework supporting the observed association between inadequate sleep and AFL recurrence.

Additional adjustment for daytime nap duration did not alter the association between nocturnal sleep and AFL recurrence, and nap duration was not independently associated with AFL recurrence in the same multivariable model. This contrasts with findings from our group in the SUN cohort, where longer habitual nap duration was associated with an increased incidence of AF in a primary prevention setting^32^. The lack of association in the present post-ablation context may indicate that, once AF has developed and structural remodelling is present, the influence of daytime sleep becomes less relevant than that of nocturnal sleep duration.

Furthermore, while inadequate nocturnal sleep duration was associated with a higher risk of AFL recurrence, no significant association was observed for AF recurrence alone. This discrepancy may reflect the possibility that sleep-related autonomic and circadian disturbances, exert a stronger influence on non-AF atrial tachyarrhythmias than on AF itself, thereby amplifying susceptibility to AFL recurrence without necessarily affecting AF recurrence^33^.

To the best of our knowledge, this is one of the few studies to examine the influence of sleep patterns on outcomes after ablation for AF. The present study has several strengths. It was conducted within the framework of a multicentre, randomized clinical trial with standardized follow-up and intensive rhythm monitoring through serial Holter recordings and weekly electrocardiograms, minimizing misclassification of arrhythmia recurrence and allowing a robust longitudinal assessment of outcomes. The cohort was large and representative of patients undergoing ablation in real-world clinical settings. In addition, the comprehensive adjustment for lifestyle and clinical factors reduces the likelihood of residual confounding, and multiple sensitivity analyses consistently supported the robustness of the observed associations.

Nonetheless, some limitations should be acknowledged. Sleep habits were self-reported rather than objectively measured, which may introduce some degree of misclassification bias. However, such measurement error is expected to be non-differential and would likely bias results toward the null. The assessment of sleep duration at baseline may not fully capture changes in sleep behaviour during follow-up, which could similarly lead to non-differential misclassification. Although the models were adjusted for a wide range of potential confounders, unmeasured factors such as sleep quality and insomnia symptoms cannot be excluded. Finally, while the study included a broad spectrum of patients undergoing ablation, the generalizability of these findings to populations with different ethnic or clinical profiles warrants further investigation.

In conclusion, this study provides novel evidence that inadequate nocturnal sleep duration (<6 h/day or >8 h/day) may be associated with an increased risk of AFL recurrence after catheter ablation for AF. These findings underscore the potential relevance of sleep habits as a modifiable factor influencing post-ablation outcomes. Incorporating the assessment of sleep duration into post-ablation follow-up may help identify patients at higher risk of arrhythmia recurrence and support more personalized lifestyle recommendations. Further research using objective sleep measurements and interventional designs is warranted to confirm these associations and elucidate the mechanisms linking sleep behaviour to the risk of arrhythmias.

## ACKNOWLEDGEMENTS

Innoliva provided the necessary amounts of extra-virgin olive oil to participants in the intervention group, and the Basque Culinary Center has collaborated in the elaboration of videos for the nutritional intervention. **The** authors thank all the volunteers of the PREDIMAR study as well as the dietitians Víctor de la O, Estíbaliz Goñi, María José Cobo, Maria Vasilj, and Ainara Martínez, the medical doctor Liz Ruiz, and the nurses and research coordinators from each recruitment center.

## AUTHORS’ CONTRIBUTIONS

JDG and MRC contributed to the conception or design of the work. JDG, MTBL, GBE, MAMG, and MRC contributed to the analysis and interpretation of data for the work. MTBL, LG, PR, LT, JLI, EC, AIC, RMR and IGB contributed to the collection and assembly of data. JDG drafted the manuscript. All critically revised the manuscript and gave final approval and agree to be accountable for all aspects of work ensuring integrity and accuracy.

## DATA AVAILABILITY

The data underlying this article will be shared on reasonable request to the corresponding author.

## Non-standard Abbreviations and Acronyms

AF: atrial fibrillation
AFL: atrial flutter
EVOO: extra-virgin olive oil
FFQ: food frequency questionnaire
LA: left atrial
MEDAS: Mediterranean Diet Adherence Screener
PREDIMAR: Prevention of Recurrent Arrhythmias with Mediterranean Diet

